# Steroidal Therapy plus Conventional Therapy versus Conventional Therapy alone for Tuberculous Meningitis: A systematic review and Meta-analysis

**DOI:** 10.1101/2024.09.05.24312952

**Authors:** Ansa Naseem, Muhammad Moiz Javed, Malaika Rehmani, Muhammad Haider Tariq, Mahnoor Sikandar, Usama Ejaz, Muhammad Shahzaib Bajwa, Tehseen Raza, Muhammad Usman Khan, Ali Shehram, Muhammad Hammad Khan, Muhammad Ayyan, Muhammad Luqman

**Author notes:** **Corresponding Author:** Muhammad Hammad Khan, MBBS; Muhammad Ayyan, MBBS, Department of Medicine, King Edward Medical University, Lahore, Pakistan Neela Gumbad, Mayo Hospital Road, Lahore, Pakistan. **Credit Author Statement: Ansa Naseem:** Conceptualization, Methodology, Formal Analysis, Software, Data curation, Writing-original draft preparation; **Muhammad Moiz Javed:** Data curation, Methodology, Investigation, Writing - original draft**; Malaika Rehmani:** Data Curation, Methodology, Investigation, Validation, Visualization, Writing - original draft; **Muhammad Haider Tariq:** Data Curation, Writing - original draft, Visualization; **Mahnoor Sikandar:** Data Curation, Writing – Original draft; **Usama Ejaz:** Data Curation, Writing – Original draft; **Muhammad Shahzaib Bajwa:** Visualization, Investigation, Validation, Writing – review & editing; **Tehseen Raza:** Investigation, Visualization, Writing – review & editing; **Muhammad Usman Khan:** Investigation, Validation, Writing – review & editing; **Ali Shehram:** Investigation, Visualization, Writing – review & editing; **Hammad Khan** Supervision, Writing – original draft, review & editing; **Muhammad Ayyan:** Supervision, Writing – review & editing; **Muhammad Luqman, MBBS^4^:** Writing – review & editing. **Declaration of Interest:** The authors have no conflict of interest. **Disclosure Statement:** The authors report no proprietary or commercial interest in any product mentioned or concept discussed in this article. **Data availability statement:** The data used in the study is available upon reasonable request to the author.

## Abstract

**Background:** Tuberculosis meningitis (TBM) is the most severe form of extrapulmonary tuberculosis with a high rate of morbidity and mortality. Treatments for TBM include conventional therapy alone as well as adjunctive use of steroidal therapy.

**Methods:** A comprehensive search of electronic databases such as PubMed, Cochrane, and Scopus was conducted from inception to April 2024 to retrieve all randomized controlled trials (RCTs) that compared steroidal therapy plus conventional antituberculosis therapy with antituberculosis therapy alone for patients of TBM. Meta-analysis was performed using Review Manager 5.4. Dichotomous outcomes were compared using risk ratio (RR).

**Results:** There was a statistically significant decrease in risk of mortality in the Steroid group compared to the control group (RR 0.84; 95% CI 0.75, 0.94; p-value < 0.05). However, there was no statistically significant difference in the risk of neurological deficit between the two groups. There was a statistically significant reduction in the risk of adverse events in the steroidal therapy group compared to the control group (RR 0.90; 95% CI 0.83, 0.98; p-value = 0.03). Similarly, there was a significant improvement in the clinical efficacy in the steroidal therapy group compared to the control (RR 1.16; 95% CI 1.02, 1.31; p-value = 0.02).

**Conclusion:** Steroids in addition to antituberculosis therapy significantly reduce mortality and adverse events, while improving clinical outcomes in patients of TBM. There is a need for controlled studies with longer follow-up durations to improve the robustness of the results.

## Introduction

Tuberculosis (TB) continues to be a major cause of mortality worldwide. When TB spreads to the brain, it leads to a severe type of extrapulmonary TB, known as tuberculous meningitis (TBM) which occurs in 1–5% of patients with TB (1). Tuberculous meningitis (TBM) had an estimated global incidence of 164,000 cases and 78,200 deaths in the adult population in 2019 (2). Tuberculous meningitis (TBM) is common in young children and HIV patients but also occurs in adults. Diagnosis and treatment are often inadequate due to non-specific symptoms that complicate diagnosis, low bacilli count in CSF causing false-negative results, and drug-resistant strains. Despite providing anti-tuberculosis treatment, TBM remains a leading cause of death and neurological damage (3).

TBM is an insidiously progressive disease, prone to rapid acceleration once neurologic deficits supervene, and often leading to death within 5 to 8 weeks (4). Early initiation of anti-tuberculosis drugs has shown effectiveness, as the clinical outcome significantly depends on recognizing and starting therapy before the onset of altered mentation or focal neurologic deficits (5). Even with conventional antituberculosis drugs, the clinical prognosis remains unfavourable in TBM. Corticosteroids are often administered alongside these drugs to alleviate meningeal swelling and reduce brain pressure, lowering the risk of death or long-term neurological impairment among those who survive (6). Although conclusive evidence on specific treatment regimen, dosage, or associated with any added benefit is still scarce, literature suggests that adding corticosteroids to anti-tuberculosis treatment improves survival in TBM patients (7).

A previous systematic review (6) addressed the benefits of corticosteroid use in treating TBM. However, it does not include recent clinical trials (8,9), one of them being the largest trial conducted on TBM. Moreover, the previous study relies considerably on studies published before the year 2000. Therefore, there is valuable new data available that has not yet been incorporated into a systematic review. Given the availability of new trials, our review aims to incorporate these recent findings and address the knowledge gap for clinical practice and further research.

## Methods

We performed this systematic review and meta-analysis according to the guidelines of the Cochrane Handbook for Systematic Reviews of Interventions and reported as per PRISMA (Preferred Reporting Items for Systematic Reviews and Meta-Analyses) (10). This review was registered under the International Prospective Register of Systematic Reviews (PROSPERO) under Prospero ID CRD42024574404. No ethical approval was required for our review.

### Search Strategy

A comprehensive search was conducted across electronic databases including PubMed, Elsevier’s Scopus, and Cochrane’s CENTRAL. Additionally, we searched international trial registers including ClinicalTrials.gov and WHO International Clinical Trials Registry Platform for literature available from inception up to April 2024. The keywords and Medical Subject Headings (MeSH) terms used included “Corticosteroids” OR “Steroidal therapy” AND “anti-tuberculous therapy” AND “Tuberculous meningitis”. A detailed search string is provided in **Supplementary File Table ST1**. The references generated by the articles that came forth were also reviewed to identify relevant studies.

### Screening of Studies

The search results were imported into the Zotero library for screening and duplicates were removed. Initial screening based on title and abstract was done by three authors. The selected studies underwent rigorous full-text screening before being included in the final review. Any discrepancies concerning study selection were resolved by a senior author.

### Eligibility criteria

Articles that met the following inclusion criteria were selected: 1. population: patients diagnosed with tuberculous meningitis and not having received prior anti-TBM medication; 2. intervention: corticosteroid therapy in addition to the anti-tuberculosis therapy; 3. comparator: anti-tuberculosis medication alone; 4. studies reporting at least one of the outcomes of interest; 5. study design: randomized controlled trials (RCTs) were included in the quantitative analysis.

Studies on solely paediatric populations and those that did not compare corticosteroid therapy in addition to anti-tuberculous therapy with anti-TB therapy directly were excluded. Review articles, case series, case reports, editorials, correspondences, and single-arm studies were excluded. Additionally, pre-prints, animal-based studies, and studies published in languages other than English were also excluded.

### Data Extraction

Data relating to the type of study, sample size, age, gender, steroid therapy regimen, and duration of follow-up were extracted from the tables, figures, and texts present in the articles by two independent investigators into a prepiloted Microsoft Excel spreadsheet. Any disparity that arose was resolved by consulting with a senior author, who rechecked the extracted data.

The outcomes were divided into primary and secondary outcomes. The primary outcomes included mortality and neurological deficit. The secondary outcomes included adverse reactions to the medication and clinical efficacy, which was defined as substantial resolution of symptoms, and/or decreased disability and dependence on others.

### Risk of bias assessment

The risk of bias in the randomized controlled trials was assessed using the revised Cochrane’s Risk of Bias tool (RoB2.0) for randomized studies (11). The tool is used to assess for risk of bias in five domains, namely: 1. bias arising from the randomization process; 2. bias caused by deviations from intended interventions; 3. bias caused by missing outcome data; 4. bias in the measurement of the outcome, and 5. bias in the selection of the reported result. Two independent reviewers assessed each study to be at low risk of bias, some concerns, or high risk of bias. A third reviewer resolved any disagreements between them.

### Statistical analysis

Statistical Analysis was conducted on Review Manager (RevMan, version 5.4; The Cochrane Collaboration, Copenhagen, Denmark) for Windows. The random effects model was used with the Mantel-Haenszel method. Dichotomous outcomes were compared using Risk Ratio (RR) and 95% confidence interval (CI). All the p-values were two-tailed, and statistical significance was set at p-value <0.05.

To assess heterogeneity, Chi-square and I^2^ statistics were used for each synthesis. The threshold for significant heterogeneity was set at a p-value < 0.1.

## Results

The search elicited a total of 1157 articles, of which ten randomized controlled trials (RCTs) were included in the meta-analysis (7–9,12–18). The study by Torok et al. was found to be a long-term follow-up of the study by Thwaites et al. The process of study screening and selection is shown in the PRISMA flowchart **Figure 1**.

**Figure 1:**
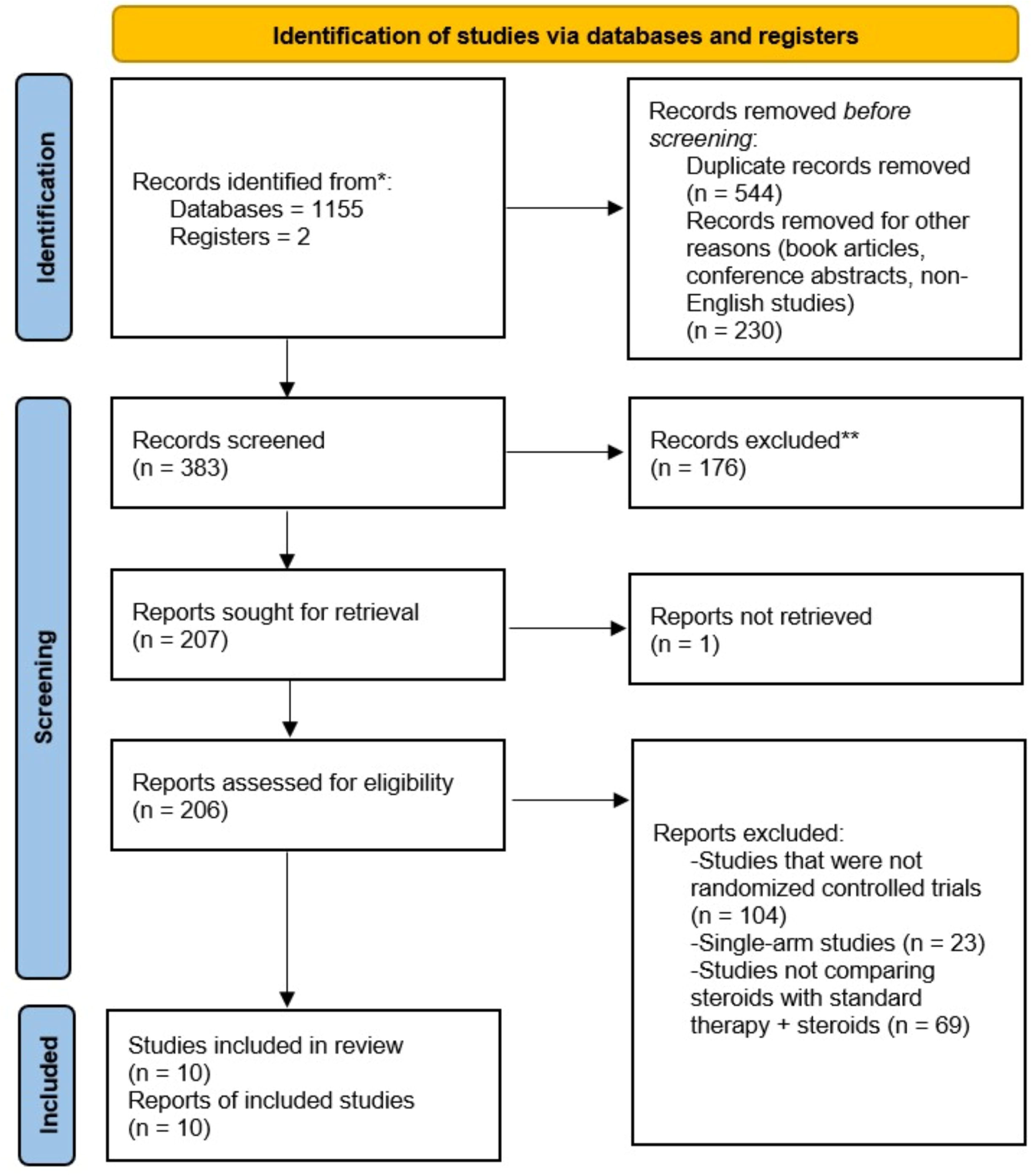
PRISMA flowchart for study screening and selection.

### Characteristics of included studies

The included RCTs reported data of 1278 patients. Two studies were double-blind, placebo-controlled trials and one was an open-label trial. In addition to the administration of anti-tuberculous drugs for the treatment of TB meningitis patients, dexamethasone was used as an adjunct therapy in seven of the RCTs. In contrast, one study used prednisolone and another one used methylprednisolone in addition to anti-tuberculosis therapy. Primary and secondary outcomes were reported at various intervals in different studies, ranging from 3 months to 5 years. Detailed baseline characteristics of the study populations can be found in **Table 1**. Further details of included studies and the intervention and control regimens are provided in the supplementary file **Table ST2 and ST3.**

**Table 1:** Study characteristics of included studies.

| **Study ID** | **Study Type** | **Location** | **Study arms**<br>**(Steroid vs**<br>**Non-**<br>**Steroid)** | **Number of**<br>**patients**<br>**randomized**<br>**(Steroid vs**<br>**Non-**<br>**Steroid)** | **Mean Age**<br>**(SD)**<br>**(Steroid vs**<br>**Non-**<br>**Steroid)** | **Male %**<br>**(Steroid**<br>**vs Non-**<br>**Steroid)** | **Duration**<br>**of**<br>**Follow-**<br>**up** |
| --- | --- | --- | --- | --- | --- | --- | --- |
| Chotmongkol 1996 | RCT | Thailand | Prednisolone vs Control | 29 vs 30 | NR | NR | 6 months |
| Diao 2020 | RCT | China | Methylprednisolone + SHREZ/6HR vs SHREZ/6HR | 34 vs 34 | 45.35±13.73 vs 45.93±13.69 | 47.1 vs 44.1 | 6 months |
| Donovan 2023 | Double-blind RCT | Indonesia and Vietnam | Dexamethasone vs Placebo | 263 vs 257 | Median: 36 (29-41) vs 36 (30-42) | 79.1 vs 73.2 | 12 months |
| Girgis 1991 | RCT | Egypt | Dexamethasone + ATT vs ATT alone | 145 vs 135 | Range: 5 months - 55 years | 55.9 vs 57.0 | 24 months |
| Green 2009 | RCT | Vietnam | ATT plus Dexamethasone vs ATT | 18 vs 19 | 25(19.3-40.5) vs 31(23.0- | 44.4 vs 36.8 | 9 months |
|  |  |  | plus Placebo |  | 44.0) |  |  |
| Kumarvelu<br>1994 | RCT | India | ATT +<br>dexamethasone Vs ATT | 24 vs 23 | 26.9 (range<br>12-78) | 53 | 3 months |
| Malhotra<br>2009 | Open<br>Label<br>RCT | India | ATT plus<br>Dexamethasone vs ATT<br>alone | 31 vs 30 | 31.97(15-66) vs<br>32.87(15-70) | 15 vs 14 | 1, 2, 6<br>and 10<br>month<br>follow up |
| O'Toole<br>1969 | RCT | India | Dexamethasone + ATT vs<br>ATT alone | 11 vs 12 | NR | NR | Weekly<br>follow-up<br>till<br>outcomes<br>were in<br>doubt |
| Thwaites<br>2004 | Double<br>Blind<br>RCT | Vietnam | ATT plus<br>dexamethasone vs ATT<br>plus placebo | 274 vs 271 | Median: 36<br>(range15-88) vs 35<br>(range 15-84) | 61.3 vs<br>60.1 | 9 months |
| Torok 2011 | Double<br>Blind<br>RCT | Vietnam | ATT plus<br>Dexamethasone vs ATT<br>plus Placebo | 274 vs 271 | Median: 36<br>(range15-<br>88) vs 35<br>(range 15- | 61.3 vs<br>60.1 | 5 years |
|  |  |  |  |  | 84) |  |  |

### Risk of bias

Upon evaluation of the risk of bias, two studies (7,13) were rated as having a low risk of bias. The remaining studies had some concerns regarding the randomization process, selection of the reported result, and measurement of the outcome by assessors who were not blinded to treatment allocation. Details of the risk of bias assessment are provided in the supplementary file **Table ST4**.

### Primary outcomes

At up to two years of follow-up, there was a statistically significant decrease in risk of mortality in the Steroid group compared to the control group (RR 0.84; 95% CI 0.75, 0.94; p-value < 0.05; I^2^=0%). However, there was no statistically significant difference in the risk of neurological deficit between the two groups (RR 0.92; 95% CI 0.66, 1.28; p-value = 0.60; I^2^=44%), shown in **Figure 2**.

**Figure 2:**
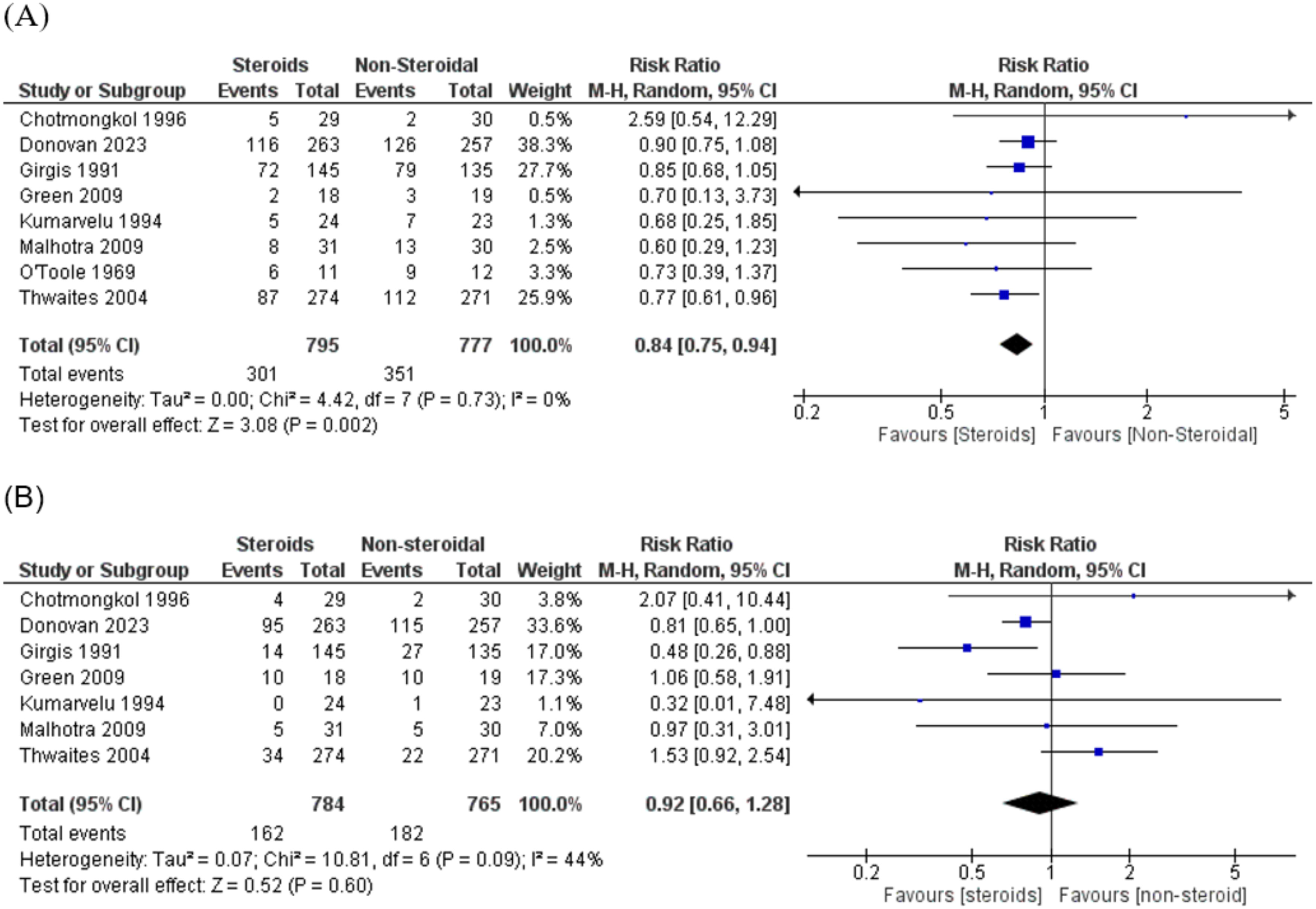
Primary outcomes at 2 years of follow-up: (A) Mortality; (B) Neurological deficit.

Only one study reported a follow-up longer than two years (18) and this was a 5-year follow-up of the study by Thwaites et al.. When study data from the longest available follow-ups from each study population was pooled, there was a statistically significant reduction in mortality risk in the steroidal therapy group (RR 0.89; 95% CI 0.80, 0.99; p-value = 0.03; I^2^=0%). Similarly, there was a significant decrease in the risk of neurological deficit in the steroidal therapy group compared to the control group (RR 0.81; 95% CI 0.68, 0.97; p-value = 0.02; I^2^=0%), shown in **Figure 3**.

**Figure 3:**
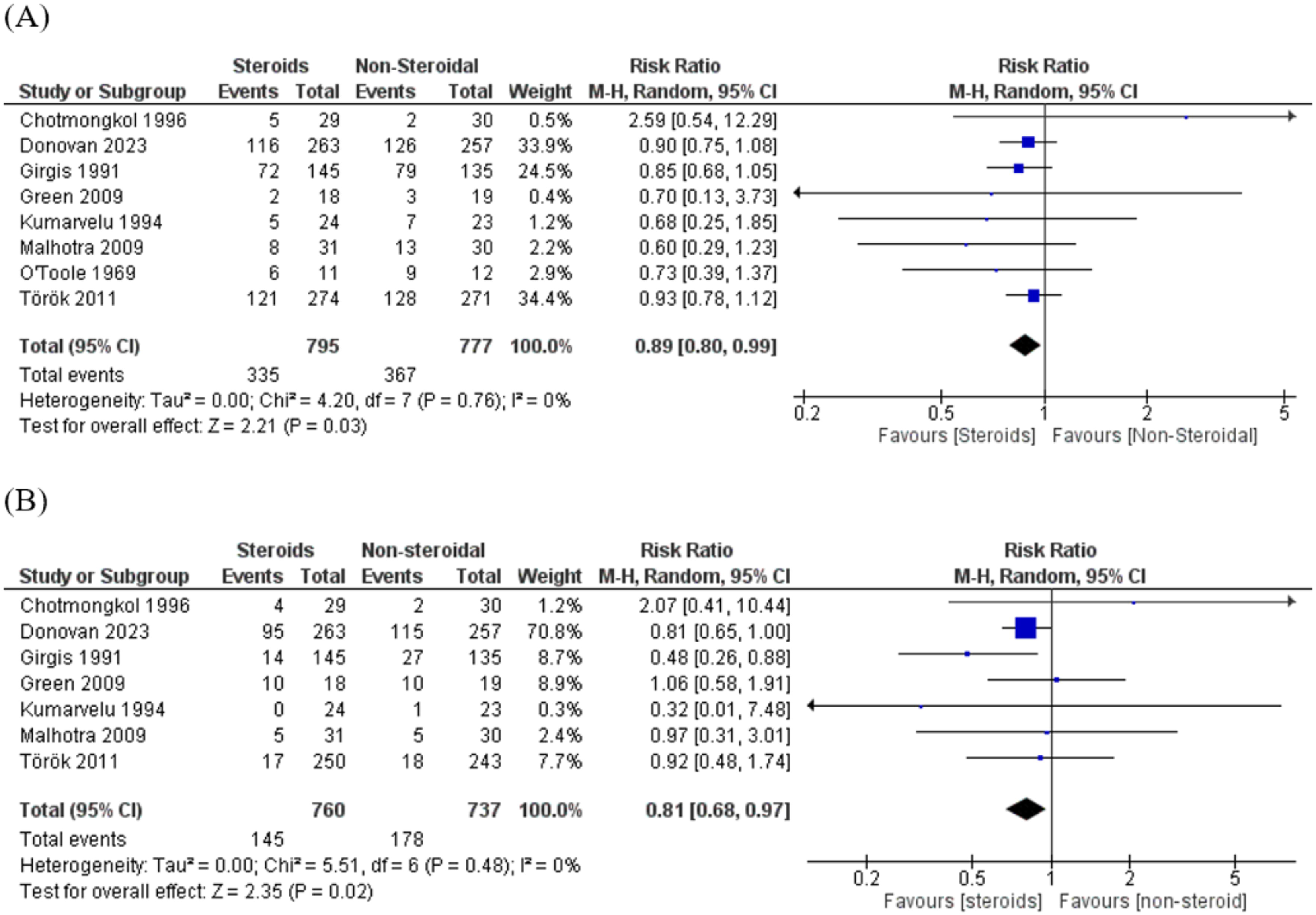
Primary outcomes at longest available follow-up: (A) Mortality; (B) Neurological deficit.

### Secondary outcomes

The secondary outcomes in our study were adverse events and clinical success in the treatment of TB meningitis. There was a statistically significant decrease in the risk of adverse events in the steroidal therapy group compared to the control group (RR 0.90; 95% CI 0.83, 0.98; p-value = 0.03; I^2^=11%). Similarly, there was a significant improvement in the clinical efficacy in the steroidal therapy group compared to the control (RR 1.16; 95% CI 1.02, 1.31; p-value = 0.02; I^2^=0%), shown in **Figure 4**.

**Figure 4:**
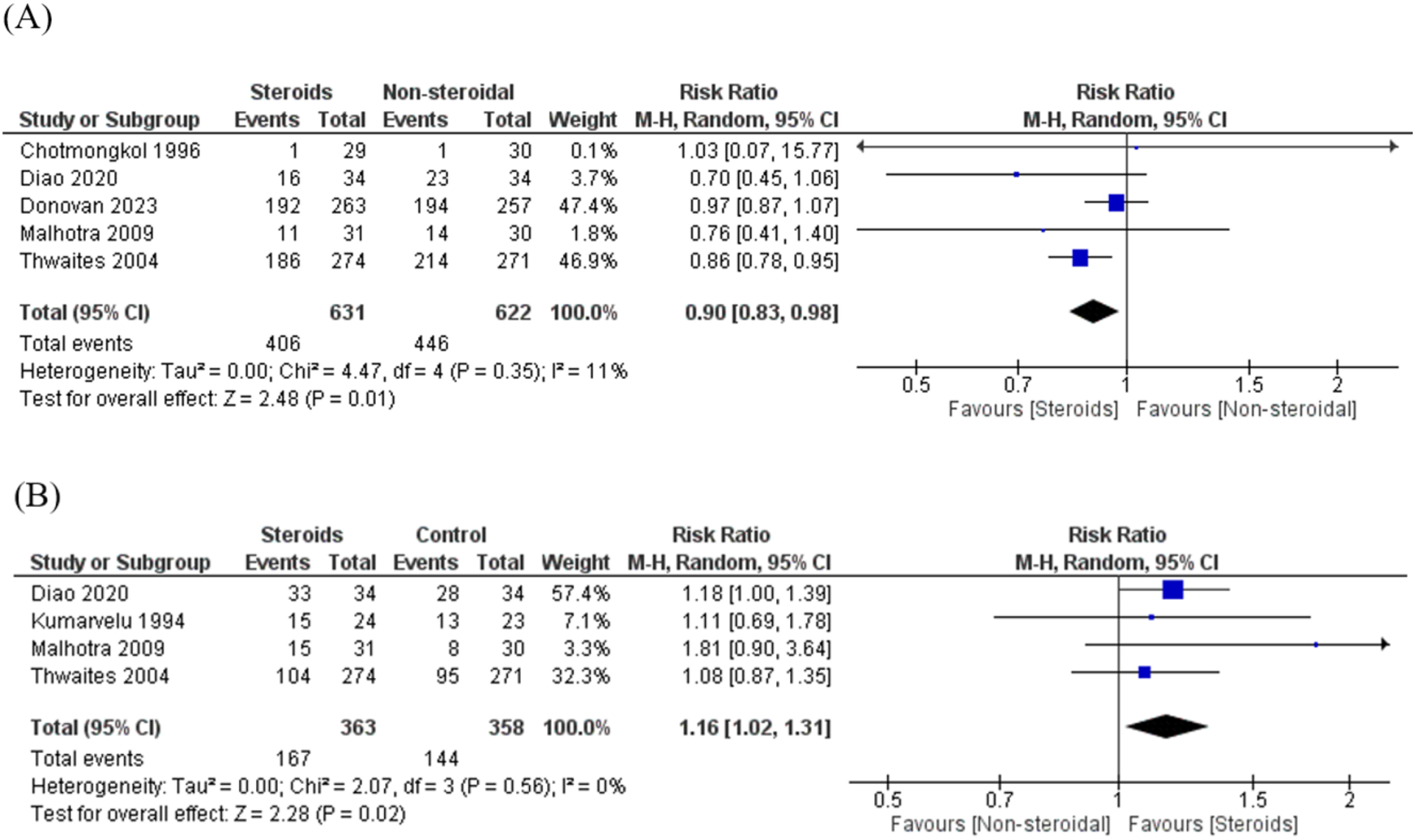
Secondary outcomes: (A) Adverse events; (B) Successful treatment of Tuberculosis meningitis.

## DISCUSSION

Our systematic review evaluated the efficacy of steroidal therapy in improving outcomes for patients with tuberculous meningitis compared to antituberculosis medication without steroids. We found a statistically significant reduction in the risk of mortality in patients treated with steroids compared to those who received the control therapy. There was an improvement in clinical success along with a decrease in adverse events in patients treated with steroids. However, there was no significant difference in the risk of neurological deficit between the two groups at comparable follow-up.

Our findings align with previous research suggesting that corticosteroids can reduce mortality in TB meningitis by mitigating the inflammatory response and reducing intracranial pressure (6). Additionally, the previous study revealed comparable neurological deficits between the two groups. In terms of adverse events, our findings of lower risk in the steroidal therapy group contrast with earlier reviews that reported no significant difference in the two groups. This variation may be attributed to the addition of significantly large patient data and potential improvements in reporting adverse events in more recently conducted newer RCTs. We also evaluated the clinical efficacy in our meta-analysis, which was taken as a significant improvement in the clinical status of the patient. There was a significant improvement seen in the steroidal therapy group compared to the control group; this was an important outcome that was not evaluated by previous reviews. This shows that patient outcomes in the steroidal therapy group are better than in the control group, with better quality of life and fewer incidences of mortality and morbidity.

Current guidelines on TBM therapy published by the WHO recommend the use of corticosteroids in addition to the standard anti-tuberculosis therapy. This recommendation is justified by lower rates of mortality, disability, and adverse events observed in patients receiving adjunct steroid therapy. Additionally, patients with greater disease severity are shown to benefit to a greater extent when receiving steroids in addition to anti-tuberculosis medication. The preferred regimen is a corticosteroid therapy (dexamethasone or prednisolone) started at TBM diagnosis and tapered over 6–8 weeks. (19) These recommendations were based on randomized controlled trials, and our meta-analysis furthers the evidence of the effectiveness of said therapy in TBM patients.

The strength of our meta-analysis lies in the inclusion of more up-to-date literature. The previous systematic review included 9 studies in the review, of which only 3 studies were published after the year 2000. With changing treatment protocols and newer drug regimens for the management of TBM, there is a need for an updated review that will help guide the treatment strategies for the near future. Moreover, one of the included studies had a solely paediatric population as the study sample (20). This differed from other studies that included patients from 12 years onwards, with the mean/median age ranging from 25 years to 46 years. To reduce heterogeneity in our pooled results, this study was excluded from our analysis. A more recent review by Wang et al. (21) did include newer studies; however, we found that a number of the studies referenced in the review were not indexed on PubMed or Google Scholar. Additionally, we could not reproduce their results as the authors of the originally included studies could not be reached. Nevertheless, the review had similar findings to our study with respect to clinical effectiveness and adverse events. However, mortality and neurological deficits were not pooled in this review.

Currently, there is still a need to evaluate the cost-effectiveness of adjunct steroid therapy in treating TBM. Steroids can potentially decrease the need for prolonged hospital stays, intensive care, and treatment of complications by lowering the risk of mortality and adverse events; research is needed to cement the possible benefits of adjunct therapy, especially in lower- and middle-income countries. Additionally, treatment therapies according to disease severity need to be explored further, as one of the included trials (7) shows promising results of altering drug doses in patients with severe infection. Since TBM is highly aggressive, patients with advanced disease may require an equally aggressive treatment strategy. An ongoing trial is currently evaluating such aggressive therapies for adult TBM (22).

### Limitations

There are a number of limitations in our review. Firstly, there is a difference in the steroid classes administered to the interventional groups (dexamethasone and methylprednisolone/prednisone) along with variations in the steroidal therapy regimen. This creates a degree of heterogeneity in the assessment of the effect of steroidal therapy. Secondly, the included studies span a wide period (1969 to 2023). Over these decades, multidrug-resistant TB has become a major concern, causing many antibiotics to be rendered less effective (23).

Simultaneously, the declining TB-related mortality rate in these decades can be attributed to public health improvements and modern medicine (24). Under these varying conditions of host, pathogen, and environment, concrete conclusions cannot be derived. Thus, there is a need to study the disease progression and management in more controlled environments. There is also a lack of long-term follow-up available for patients of TBM treated with steroids in the literature. We came across only a single study reporting data at 5 years, and further studies are needed with similarly long-term follow-ups to ascertain the long-term benefits of steroidal therapy in TBM.

### Conclusion

There is a significant improvement in the rate of clinical success, with lower mortality and adverse event risk in patients of tuberculous meningitis treated with steroidal therapy in addition to the standard anti-tuberculosis treatment. There is a need for studies with more homogenous populations and longer-term follow-up to improve the robustness of these results. Additionally, a cost-benefit analysis comparing steroidal therapy with non-steroidal therapy and other treatment strategies shall allow for the adoption of the most appropriate treatment in each setting.

## Data Availability

Data is available upon reasonable request to the authors.

## Notes

**Funding:** The authors received no funding.

### Competing Interest Statement

The authors have declared no competing interest.

### Funding Statement

The study received no funding.

