## Supplementary File 1 for "Steroidal Therapy plus Conventional Therapy versus Conventional Therapy alone for Tuberculous Meningitis: A systematic review and Meta-analysis"

| **Database** | **Search Strategy** | **Results** |
| --- | --- | --- |
| **PubMED** | ("Tuberculous meningitis" OR "Tuberculosis meningitis" OR "TB meningitis") AND ("Steroidal therapy" OR "Corticosteroid" OR "Dexamethasone" OR "methylprednisone") | 165 |
| **Scopus** | ("Tuberculous meningitis" OR "Tuberculosis meningitis" OR "TB meningitis") AND ("Steroidal therapy" OR "Corticosteroid" OR "Dexamethasone" OR "methylprednisone") | 919 |
| **Cochrane CENTRAL** | (("Tuberculous meningitis" OR "Tuberculosis meningitis" OR "TB meningitis") AND ("Steroidal therapy" OR "Corticosteroid" OR "Dexamethasone" OR "methylprednisone")):ti,ab,kw | 71 |

**ST1: Detailed search strategy and search results**

**ST2: Details of study characteristics**

| **Study ID** | **Study Type** | **Location** | **Study arms (Steroid vs Non-Steroid)** | **Number of patients randomized (Steroid vs Non-Steroid)** | **Age Inclusion Criteria** | **Mean Age (SD)**  **(Steroid vs Non-Steroid)** | **Male % (Steroid vs Non-Steroid)** | **Comorbidities** | **Duration of Follow-up** |
| --- | --- | --- | --- | --- | --- | --- | --- | --- | --- |
| Chotmongkol 1996 | RCT parallel group | Thailand | Prednisolone vs Control | 29 vs 30 | >15 years | NR | NR | HIV excluded | 6 months |
| Diao 2020 | RCT |  | Methylprednisolone + SHREZ/6HR vs SHREZ/6HR | 34 vs 34 | > 18 years | 45.35±13.73 vs 45.93±13.69 | 47.1 vs 44.1 | NR | 6 months |
| Donovan 2023 | Double-blind RCT |  | Dexamethasone vs Placebo | 263 vs 257 | > 18 years | Median: 36 (29-41) vs 36 (30-42) | 79.1 vs 73.2 | HIV positive | 12 months |
| Girgis 1991 | RCT | Egypt | Dexamethasone + ATT vs ATT alone | 145 vs 135 | All | Range: 5 months - 55 years | 55.9 vs 57.0 | NR | 24 months |
| Green 2009 | RCT |  | ATT plus Dexamethasone vs ATT plus Placebo | 18 vs 19 | >14 years | 25(19.3-40.5) vs 31(23.0-44.0) | 44.4 vs 36.8 | NR | 9 months |
| Kumarvelu 1994 | RCT | India | ATT + dexamethasone Vs ATT | 24 vs 23 | >12 years | 26.9 (range 12-78) | 53 | NR | 3 months |
| Malhotra 2009 | Open Label RCT | India | ATT plus Dexmethasone vs ATT alone | 31 vs 30 | >14 years | 31.97(15-66) vs 32.87(15-70) | 15 vs 14 | NR | 1, 2, 6 and 10 month follow up |
| O'Toole 1969 | RCT | India | Dexamethasone + ATT vs ATT alone | 11 vs 12 | All | NR | NR | NR | Weekly follow-up till outcomes were in doubt |
| Thwaites 2004 | Double Blind RCT | Vietnam | ATT plus dexamethasone vs ATT plus placebo | 274 vs 271 | >14 years | Median: 36 (range15-88) vs 35 (range 15-84) | 61.3 vs 60.1 | NR | 9 months |
| Torok 2011 | Double Blind RCT | Vietnam | ATT plus Dexamethasone vs ATT plus Placebo | 274 vs 271 | >14 years | Median: 36 (range15-88) vs 35 (range 15-84) | 61.3 vs 60.1 | NR | 5 years |

**ST3: Details of intervention and control therapy**

| **Study ID** | **Intervention Regimen** | **Control Regimen** |
| --- | --- | --- |
| Chotmongkol 1996 | **Prednisolone + ATT**  Prednisolone orally on tapering dosage for 5 weeks | **ATT**: isoniazid oral (300 mg), rifampicin oral (600 mg, 450 mg for those weighing < 50 kg), pyrazinamide oral (1500 mg), and streptomycin intramuscular (750 mg) for the first 2 months; followed by isoniazid and rifampicin in above dosage for 4 months. |
| Diao 2020 | **Methylprednisolone + SHREZ**  Methylprednisolone (0.5 g), intravenous drip, 500 mg/d, was administered. After 7 days of impact treatment, it was changed to 80 mg/d, and according to the patient’s condition, the dosage was reduced until a later period of oral maintenance dose treatment for a total of 3 months. | **SHREZ** for one month [that included S=Streptomycin (0.75g/time, time/day intramuscular), H=isoniazid (0.3g/time, time/day orally with empty stomach), R=rifampin (0.45g/time, time/day orally with empty stomach), E=ethambutol (0.75g/time, time/day orally with empty stomach) and Z=pyrazinamide (1.5g/time, time/day)] followed by HR only for 6 months |
| Donovan 2023 | **Dexamethasone + ATT**  **Participants with grade I disease:** intravenous dexamethasone administration for 3 weeks (0.3 mg per kilogram per day for the first week, 0.2 mg per kilogram per day for the second week, and 0.1 mg per kilogram per day for the third week) and then oral administration for 3 weeks, starting at 3 mg per day and decreasing by 1 mg each week.  **Participants with grade II or III disease:** intravenous dexamethasone administration for 4 weeks (0.4 mg per kilogram of body weight per day for the first week, 0.3 mg per kilogram per day for the second week, 0.2 mg per kilogram per day for the third week, and 0.1 mg per kilogram per day for the fourth week) and then oral administration for 4 weeks, starting at 4 mg per day and decreasing by 1 mg each week. | **ATT**: For previously untreated participants: oral isoniazid (5 mg/kg), rifampicin (10 mg/kg), pyrazinamide (25 mg/kg, maximum, 2 g/day), and intramuscular streptomycin (20 mg/kg, maximum 1 g/day) for 3 months followed by 6 months of isoniazid, rifampicin, and pyrazinamide at the same daily doses.  Ethambutol (20 mg/kg; maximum 1.2/day) was substituted for streptomycin in HIV-positive participants and was added to the regimen for 3 months for participants previously treated for TB. |
| Girgis 1991 | **Dexamethasone + ATT**  Dexamethasone given intramuscularly (12 mg/day to adults and 8 mg/day to children weighing < 25 kg) for 3 weeks and then tapered during the next 3 weeks). | **ATT**: isoniazid (10 mg/kg/day, maximum 600 mg) intramuscularly for 2 weeks then orally for 2 years, streptomycin intramuscular (25 mg/kg/day, maximum 1000 mg) for 6 weeks, and ethambutol oral (25 mg/kg/day, maximum 1200 mg) for 6 weeks, then 15 mg/kg/day for 2 years. |
| Green 2009 | **Dexamethasone sodium phosphate + ATT**  **Dexamethasone** dose based on disease severity:  Grade I disease: intravenous dexamethasone sodium phosphate 0.3 mg/kg/day in 1st week and 0.2 mg/kg/day in 2nd week followed by 4 weeks of oral dexamethasone (0.1 mg/kg/day for week 3 then a total of 3 mg/day, decreasing by 1 mg each week per week until zero).  Grade II and III disease: intravenous dexamethasone sodium phosphate given 0.4 mg/kg/day in 1st week , 0.3 mg/kg/d in 2nd week , 0.2 mg/kg/d in 3rd week , and 0.1 mg/kg/day in 4th week , and then oral dexamethasone for 4 weeks decreasing by 1 mg each week per week until zero. | **ATT:** oral isoniazid (5 mg per kilogram of body weight), rifampin (10 mg per kilogram), pyrazinamide (25 mg per kilogram; maximum, 2 g per day), and intramuscular streptomycin (20 mg per kilogram; maximum, 1 g per day) all daily for three months, followed by isoniazid, rifampin, and pyrazinamide at the same daily doses for six months. |
| Kumarvelu 1994 | **Dexamethasone** **+ ATT**  **Dexamethasone:** Intravenous 16mg/day in 4 divided doses for 7 days, then oral tablet 8 mg/day for 21 doses, and in children 0.6mg/kg/day for 7 days, reducing to 0.3 mg/kg/day for 21 days. | **ATT:** rifampicin (450 mg), isoniazid (300 mg), and pyrazinamide (1500 mg) all oral daily;  For those weighing <30 kg: 15mg/kg rifampicin, 10mg/kg isoniazid, and 30mg/kg pyrazinamide. |
| Malhotra 2009 | **Dexamethasone** **+ ATT**  **Dexamethasone** group was given IV dexamethasone for 4 weeks at 0.4, 0.3, 0.2, and 0.1 mg/kg/day during weeks 1,2,3,4 respectively followed by daily oral doses of 4,3,2,1 mg/day for weeks 5,6,7,8 respectively. | **ATT** was given in both groups as isoniazid (10mg/kg/day), rifampin (15mg/kg/day), pyrazinamide (30mg/kg/day), and ethambutol(20mg/kg/day) or streptomycin (15mg/kg/day) for 2 months and then just isoniazid (10mg/kg/day) and rifampin(15mg/kg/day) for 7 months. |
| O'Toole 1969 | **Dexamethasone** **+ ATT**  **Dexamethasone** given for up to 4 weeks in an adult dose of 9 mg/day during the first week, 6 mg/day during the second week, 3 mg/day during the third week, and 1.5 mg/day during the 4th week. | **ATT**: isoniazid intramuscular or oral (10 mg/kg/day, except in children < 2 years of age who received 20 mg/kg/day) and streptomycin (20 mg/kg/day, maximum 1 g). |
| Thwaites 2004  And  Torok 2011 | **Dexamethasone** + **ATT**  **Dexamethasone** given by disease severity: Grade I disease: intravenous dexamethasone sodium phosphate 0.3 mg/kg/day for week 1 and 0.2 mg/kg/day for week 2 followed by 4 weeks of oral dexamethasone (0.1 mg/kg/day for week 3 then a total of 3 mg/day, decreasing by 1 mg each week).  Grade II and III disease: intravenous dexamethasone sodium phosphate given 0.4 mg/kg/day for week 1, 0.3 mg/kg/d for week 2, 0.2 mg/kg/d for week 3, and 0.1 mg/kg/day for week 4, and then oral dexamethasone for 4 weeks decreasing by 1 mg each week. | **ATT**: For previously untreated participants: oral isoniazid (5 mg/kg), rifampicin (10 mg/kg), pyrazinamide (25 mg/kg, maximum, 2 g/day), and intramuscular streptomycin (20 mg/kg, maximum 1 g/day) for 3 months followed by 6 months of isoniazid, rifampicin, and pyrazinamide at the same daily doses; ethambutol (20 mg/kg; maximum 1.2/day) substituted for streptomycin in HIV-positive participants and was added to the regimen for 3 months for participants previously treated for TB. |

**ST4: Risk of Bias of included studies.**

| **Study ID** | **Randomisation process** | **Deviations from the intended interventions** | **Missing outcome data** | **Measurement of the outcome** | **Selection of the reported result** | **Overall** |
| --- | --- | --- | --- | --- | --- | --- |
| **Diao 2020** | 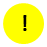 | 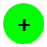 | 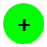 | 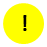 | 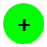 | 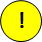 |
| **Donovan 2023** | 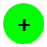 | 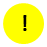 | 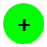 | 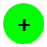 | 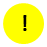 | 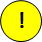 |
| **Kumarvelu 1994** | 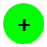 | 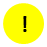 | 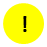 | 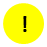 | 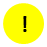 | 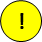 |
| **Green 2009** | 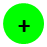 | 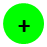 | 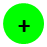 | 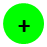 | 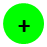 | 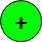 |
| **Chotmongkol 1996** | 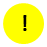 | 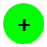 | 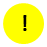 | 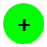 | 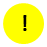 | 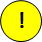 |
| **Girgis 1991** | 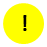 | 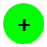 | 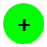 | 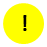 | 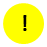 | 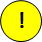 |
| **Malhotra 2009** | 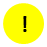 | 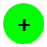 | 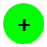 | 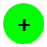 | 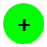 | 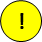 |
| **O'Toole 1969** | 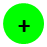 | 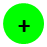 | 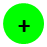 | 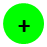 | 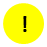 | 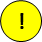 |
| **Thwaites 2004** | 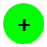 | 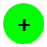 | 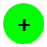 | 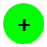 | 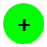 | 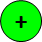 |

|  | **Low risk** |
| --- | --- |
|  | **Some concerns** |
|  | **High risk** |
